# Comparative Evaluation of CHROMagar COL-*APSE*, MicroScan Walkaway, ComASP Colistin, and Colistin MAC Test in Detecting Colistin-resistant Gram-Negative Bacteria

**DOI:** 10.1101/2019.12.17.19015156

**Authors:** John Osei Sekyere, Arnold Karabo Sephofane, Nontombi Mbelle

**Author notes:** **Address correspondence** to John Osei Sekyere.

## Abstract

Colistin has become a critical antibiotic for fatal Gram-negative infections owing to the proliferation of multidrug-resistant carbapenemase-producing bacteria. Thus, cheaper, faster, efficient and easier-to-use colistin diagnostics are required for clinical surveillance, diagnoses and therapeutics. The sensitivity, specificity, major error (ME), very major error (VME), categorial agreement, essential agreement, turnaround time (TAT), average cost, and required skill for four colistin resistance diagnostics viz., CHROMagar COL-*APSE*, ComASP Colistin, MicroScan, and Colistin MAC Test (CMT) were evaluated against broth microdilution (BMD) using 84 Gram-negative bacterial isolates. A multiplex PCR (M-PCR) was used to screen all isolates to detect the presence of the *mcr-1* to *mcr-5* genes. A 15-point grading scale was used to grade the tests under skill, ease, processing time etc. *mcr-1* was detected by both M-PCR and CMT in a single *E. coli* isolate, with other PCR amplicons suggestive of *mcr-2, -3* and *-4* genes being also observed on the gel. The sensitivity and specificity of CHROMagar COL-*APSE*, MicroScan, and ComASP Colistin, were 82.05% and 66.67%, 92.31% and 76.92%, and 100% and 88.89% respectively. The MicroScan was the most expensive at a cost (per sampe tested) of R221.6 ($15.0), followed by CHROMagar COL-*APSE* (R118.3; $8.0), M-PCR (R75.1; $5.1), CMT (R20.1; $1.4) and ComASP Colistin (R2.64; $0.2). CHROMagar was the easiest to perform, followed by ComASP Colistin, M-PCR, MicroScan, CMT and BMD whilst M-PCR and MicroScan required higher skill. The ComASP Colistin was the best performing diagnostic test, with low VME and ME, making it recommendable for routine colistin sensitivity testing in clinical laboratories; particularly, in poorer settings. It is however limited by a TAT of 18-24 hours.

**Highlights:**

- The diagnostic efficiencies, cost, required skill and ease-of-use of colistin-resistance diagnostics were evaluated against broth microdilution
- The ComASP was most sensitive, less skill-requiring and cheap
- The CHROMagar COL-*APSE* was easiest to perform, albeit expensive and less sensitive
- The MicroScan was most expensive, requires advanced skills and was very sensitive, particularly for Enterobacteriaceae without *E. cloacae* and *Salmonella*.
- The Colistin MAC test is a better PCR alternative for detecting *mcr-1* producers in low-resource settings

## Introduction

Subsequent to the exponential dissemination of extended-spectrum β-lactamases (ESBLs) in Gram-negative bacteria (GNB) and the increased use of carbapenems to treat infections unresponsive to the penicillins and cephems (cephalosporins and cephamycins), carbapenemase-producing and carbapenem-resistant GNB are becoming common worlwide ^1,2^. Thus, carbapenems, which are last-resort antibiotics, are being replaced by colistin, a polymyxin that was discovered many years ago but shelved over nephrotoxicity and neurotoxicity concerns ^2,3^. Resistance to colistin, due to its increased use to treat carbapenem-resistant GNB, has also emerged ^2–4^. Beginning from 2016, colistin resistance, which hitherto was mediated my chromosomal mutations, was found to be also mediated by plasmid-borne *mcr* genes ^5,6^. The high fatalities caused by colistin and/or carbapenem-resistant GNB infections have led to their being categorised as high-priority pathogens by the WHO ^7,8^. The clinical importance of colistin-resistant GNB (CR-GNB) necessitates an efficient diagnostic protocol for quickly and efficiently detecting them to pre-empt further escalation and mortalities ^9^.

Several diagnostic assays have been developed to detect CR-GNB, although the CLSI and EUCAST recommend the broth microdilution (BMD) as the standard test for confirming colistin sensitivity ^9^. To date, settling on an efficient, fast, cheaper and easy-to-use diagnostic for detecting colistin resistance in routine microbiology laboratories remains a challenge, particularly in developing countries ^9–11^. This is inspite of the fact that several colistin-resistance diagnostics exist, broadly categorised under phenotypic and molecular tests ^9^. The phenotypic tests include the culture-based screening media such as CHROMagar COL-*APSE*, LBJMR, Superpolymyxin etc., MIC (minimum-inhibitory concentration)-determiners such as BMD, E-test, Sensititre, ComASP Colistin, MicroScan, BD-Phoenix, UMIC, Micronaut-S, and Vitek, chelator-based assays etc. ^9^

We are still behind in the race against antimicrobial resistance and current diagnostics still have a lot of optimisation and evaluation to be done. This is particularly so due to the ever-evolving nature of antimicrobial resistance mechanisms. Owing to the dearth of evaluation studies comparing assays from screening media, MIC-determiners, chelator-based tests, and molecular assays as well as the absence of studies describing the cost and skill component of these tests, this study was undertaken ^9^. Due to logistical and reagent availability restrictions and delays, we were only able to compare the following tests selected from the various subsets of colistin diagnostics: CHROMagar COL-*APSE* (screening media), MicroScan Walkaway system (automated MIC-determiner), ComASP Colistin (manual MIC-determiner; formerly SensiTest Colistin), colistin-MAC test (chelator-based test) and multiplex PCR (molecular assay). By including a cost and required-skill component, we aim to provide data for informing colistin resistance diagnostic choices for under-resourced laboratories.

## Methods

### Bacterial sampling and colistin sensitivity testing

A total of 84 clinical GNB isolates were used for this study. The isolates were obtained from the National Health Laboratory Services (NHLS), Pretoria, and Ampath laboratories (Johannesburg) with already de-identified patient data. The species of the isolates were determined using the MicroScan Walkaway ID panels (Beckman Coulter, USA) according to the manufacturer’s instructions.

#### Determining MIC with broth microdilution

The isolates were screened with BMD using recommended CLSI (Clinical and Laboratory Standards Institute) guidelines ^12^ to identify isolates that were resistant and susceptible to colistin. Reference broth microdilution was performed in 96-well trays as briefly described below. The manufacturer’s instructions were followed for reconstituting the colistin sulphate powder. The amount of powder to be used was determined using the following formula:

Mass(mg)=[Volume(mL)×Concentration(µg/mL)]/Potency(µg/mg). To convert the potency of colistin expressed in units/mg to µg/mL, the activity expressed in units/mg was divided by 30 units/mg. The volume(mL) of diluent required was determined using the following formula: Volume(mL)=[Mass(mg)×Potency(µg/mg)]/Concentration(µg/mL). A volume of 15mL and concentration of 1280 µg/mL was used. Sterile water was used as a solvent and diluent as per CLSI guidelines.

Cation-adjusted Mueller-Hinton broth was used to prepare the appropriate working solution. The volume (mL) of stock solution needed was determined using the following formula:

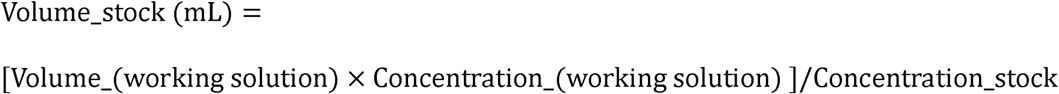

The volume required for the working solution depended on the number of test samples and the number of dilutions required. A volume of 1mL of 1280 µg/mL of stock solution was diluted with 9 mL of cation-adjusted Mueller-Hinton broth to make a working solution with a concentration of 128µg/mL and a volume of 10mL. After preparing the working solution, 100mL of the broth medium was added to each well of the microtiter plate. A volume of 100µL of the colistin working solution (128µg/mL) was added to the first well of each row on the microtiter plate. The latter resulted in a 1:2 dilution of the working solution and was the starting (highest) concentration tested. A multichannel pipette was used and set to a volume of 100 µL and the colistin broth mixture was mixed in well 1. The 100 µL colistin-broth mixture was transferred from well 1 to 2 and mixed. The process was repeated until well 10. After mixing of well 10, the 100 µL in the pipette is discarded. Wells 11 and 12 only contained the growth medium. The prepared microtiter plates were used immediately for antimicrobial susceptibility testing and those not used were stored in a -80°C refrigerator.

The direct colony suspension method was used. Fresh cultures of each test organism were prepared on blood agar plates and incubated overnight. Isolated colonies from the fresh cultures was used to prepare a saline suspension of the organism equivalent to a 0.5 McFarland turbidity standard. The 0.5 McFarland standard was measured using a spectrophotometer to ensure accuracy. Within 15 minutes of preparation, 0.3 mL of the 0.5 McFarland turbidity standard bacterial suspension was added to 2.7 mL of sterile saline. This resulted in a 1:10 dilution and yielded a concentration of 10^7 CFU/mL. A final solution of 5×10^5CFU/mL in each well was prepared by inoculating 5 µL of the 10^7 CFU/mL suspension into a well of the microtiter plate containing 100 µL volume.

The prepared microtiter trays and standardised and adjusted bacterial suspension were used. Within 15 minutes of inoculum preparation, wells 1 to 10 and 12 were inoculated with 5 µL of the inoculum. Well 11 served as the negative control and well 12 served as the positive (growth) control with broth only. Eight isolates were inoculated into a single 96 well tray. An aliquot was taken from well 12 and inoculated on blood agar and incubated overnight according to Table 2 below to ensure purity. After all wells were inoculated, the tray was agitated at 240 revolutions per minute for at least 1 minute to ensure even distribution of the inoculum. The trays were stacked four high and sealed in plastic bags to prevent drying prior to incubation. After incubation, the microtiter trays were removed from the incubator and placed on an agitator at 240 revolutions per minute for at least 1 minute to ensure uniform dispersal of any growth. The microtiter trays were placed on a reading grid and MIC values were read by observing growth in each well macroscopically by turbidity in the well. All tests were carried out in duplicate and a third test was carried out and a modal value was considered when there were discordant results between tests. *Pseudomonas aeruginosa* ATCC 27853 and *Escherichia coli* ATCC 25922 Quality Controls strains were used.

The EUCAST breakpoints (susceptible ≤ 2 mg/L; resistant > 2 mg/L) were used to interpret the results due to an absence of CLSI breakpoint for colistin ^9,13^.

### Detection of mcr-producing bacteria with multiplex polymerase chain reaction (PCR)

Genomic DNA was isolated from overnight cultures of the isolates using the Zymo Research Quick-DNA™ miniprep Kit (USA) according to the manufacturer’s instructions. A multiplex PCR (M-PCR) assay was used to determine the presence or absence of mobile colistin resistance genes, *mcr-1* to *mcr-5*, using already described protocols and primers (Table 1) ^9,14^. Primers were procured from Inqaba biotechnologies (Pretoria, South Africa).

**Table 1.**
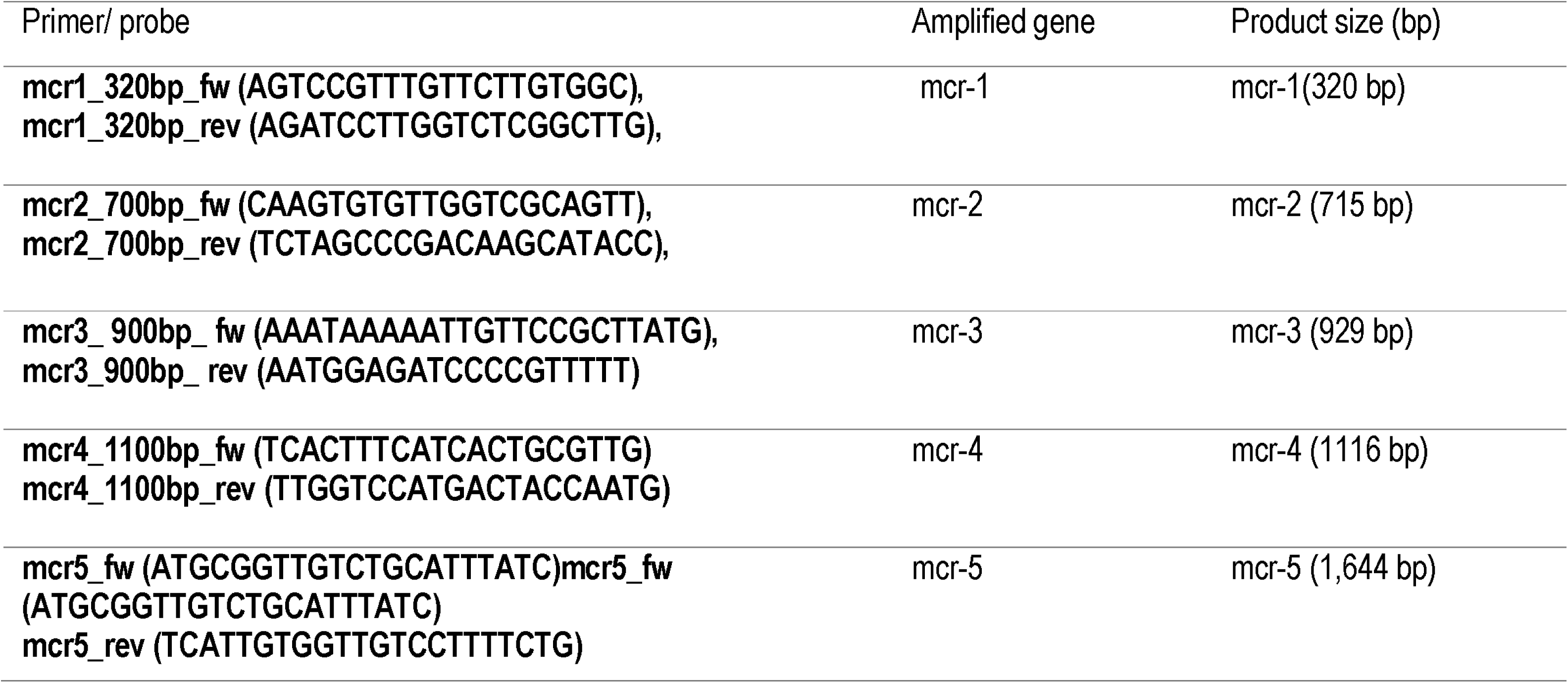
Primers used in real-time multiplex PCR to detect the mcr-1 to mcr-5 genes.

**Table 2.** Colistin BMD plates Incubation conditions.

| Organism | Incubation conditions | Incubation time |
| --- | --- | --- |
| *Enterobacteriaceae*, Non-<br>*Enterobacteriaceae* and<br>*Pseudomonas aeruginosa* | 35 +/-; Ambient air | 16-20 hours |
| *Acinetobacter* spp. | 35 +/- Ambient | 16-20 hours |

The PCR cycling conditions for the multiplex PCR were carried out as illustrated in a review by Osei Sekyere *et al*. (2018). The first cycle of denaturation was performed using a thermocycler (Bio-rad T100™ Thermal cycler, Bio-rad Laboratories Inc, USA) at 94°C for 15 min, followed by 25 denaturation cycles at 94°C for 30 seconds and annealing set at 58°C for 90 seconds. Elongation was done at 72°C for 60 seconds and the final elongation cycle at 72°C for 10 min. The amplified products were visualised using a visualisation software (DigiDoc-It 120, UVP, LCC, USA) on a 1.5% agarose gel at a voltage of 130V. Ethidium bromide (Sigma-Aldrich, USA) was used as an intercalating agent.

### *Identifying colistin-resistant isolates with CHROMagar COL-APSE, MicroScan Walkaway system, ComASP Colistin* and Colistin MAC Test (CMT)

Fresh 24-hour cultures of the isolates, grown on blood agar (Oxoid, UK), were plated unto already prepared CHROMagar™ COL-*APSE* media (ChromAgar, Paris, France) using manufacturer’s instructions. The inoculated plates were incubated at 37°C under aerobic conditions overnight and the results were observed by observing different coloured colonies, using the manufacturer’s interpretation criteria for identifying CR-GNB. The media’s ability to identify and correctly differentiate CR-GNB species within mixed cultures was assessed and compared to the MicroScan Walkaway system’s species identification.

For the MicroScan analyses, a single colony from the overnight cultures on the blood agar was standardized promptly using the prompt inoculation system provided in the reagent packaging for MicroScan AST (antimicrobial sensitivity testing) and identification (ID) testing. After inoculation, the solution was transferred into the N66 panels and loaded into the instrument for overnight processing. Species identification was obtained and colistin sensitivity results were interpreted according EUCAST breakpoints (susceptible ≤ 2 mg/L; resistant > 2 mg/L).

ComASP COL (colistin) consists of a compact panel of four tests containing seven two-fold dilutions of dehydrated COL. In this study, a 0.5 McFarland suspension of the isolates was prepared in a solution of 250mL saline and then diluted to 1:20 in saline (Gibco, Thermo Fisher Scientific, Massachusetts, USA) to obtain the first solution (Solution A). Solution A (0.4mL) was added to a Mueller-Hinton broth (3.6mL) tube to obtain solution B. One hundred microliters of solution B was dispensed into each well and the panels were incubated (Vacutec, US) at 36±2°C for 16 to 20 hours in ambient air. Results were analysed visually using a bright indirect light against a dark background.

The CMT was carried out as described in a study by Coppi *et al*. (2018)^15^. The colistin sulphate concentrations tested ranged from 0.125 to 64µg/mL alone or in combination with DPA (dipicolinic acid) at a fixed concentration of 900 µg/mL. The DPA stock solution was prepared in dimethyl sulphoxide (DMSO) at a concentration of 100mg/mL and stored at - 20°C. Cation-adjusted Mueller-Hinton broth was used as the medium for susceptibility testing alongside DPA and DMSO at a fixed concentration of 900 µg/mL and increasing colistin concentrations (0.125-64µg/mL). Wells 11 and 12 were used for growth control whilst well 11 was not inoculated; well 12 was inoculated with the test isolate. Susceptibility testing was carried out in 96-well microtitre trays with results recorded after 20 hours of incubation at 35 ± 2°C. Susceptibility to colistin was interpreted using the EUCAST guidelines: Resistant > 2mg/L; Susceptible ≤ 2mg/L. The results were considered as *mcr*-1 positive (a ≥8-fold reduction) or *mcr*-1 negative (a ≤2-fold reduction) in the presence of DPA.

### Data and statistical analysis

The results of the various tests were curated in Microsoft Excel and categorised into susceptible and resistant isolates using the BMD results. Using the BMD as the gold standard, the sensitivity, specificity, positive and negative predictive values (PPV and NPV respectively), categorical agreement (CA), essential agreement (EA), major errors (ME) and very major errors (VME) of each test were calculated using the equations 1-8 below.

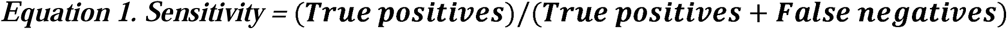

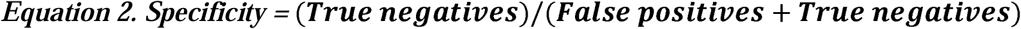

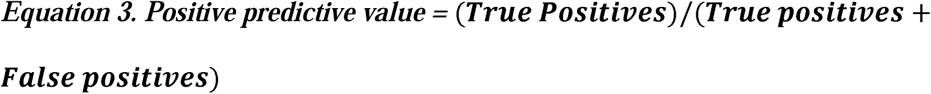

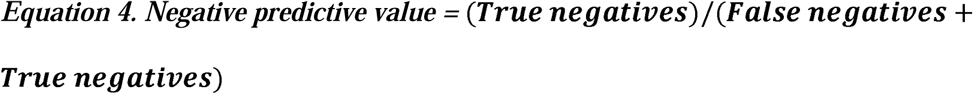

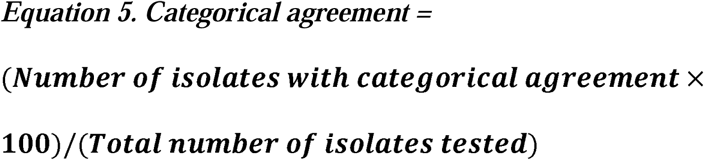

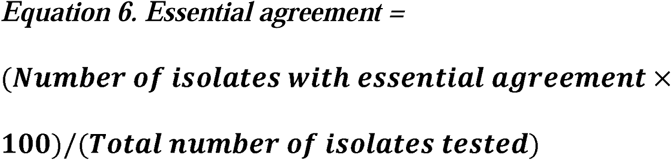

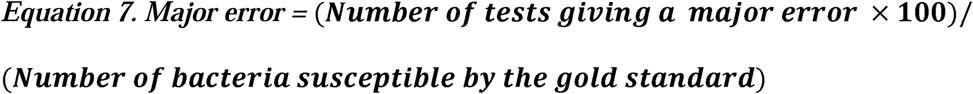

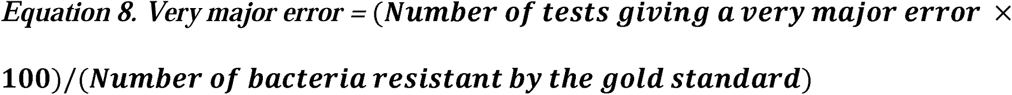

### Cost and skill (ease of application) analyses

The cost per sample for each of the tests were calculated by dividing the total cost of reagents, including VAT, by the number of samples that were tested by those reagents. Equipment costs, specifically the MicroScan Walkaway system and the thermocycler used for the M-PCR, were not included. The costs were calculated in Rands and converted to US dollars using current exchange rates (12-13/12/2019).

We assessed the required skill and ease of using the various tests by grading the various tests against five parameters: (1) time in hours needed to perform each test; (2) ease in carrying out test; (3) skill or training required (4) requirement for 3^rd^-party equipment and infrastructure and (5) cost and access to 3^rd^-party equipment and infrastructure. The breakdown of the assigned grades, on a scale of 5 to 15 for these five parameters, are shown in Table 3.

**Table 3.**
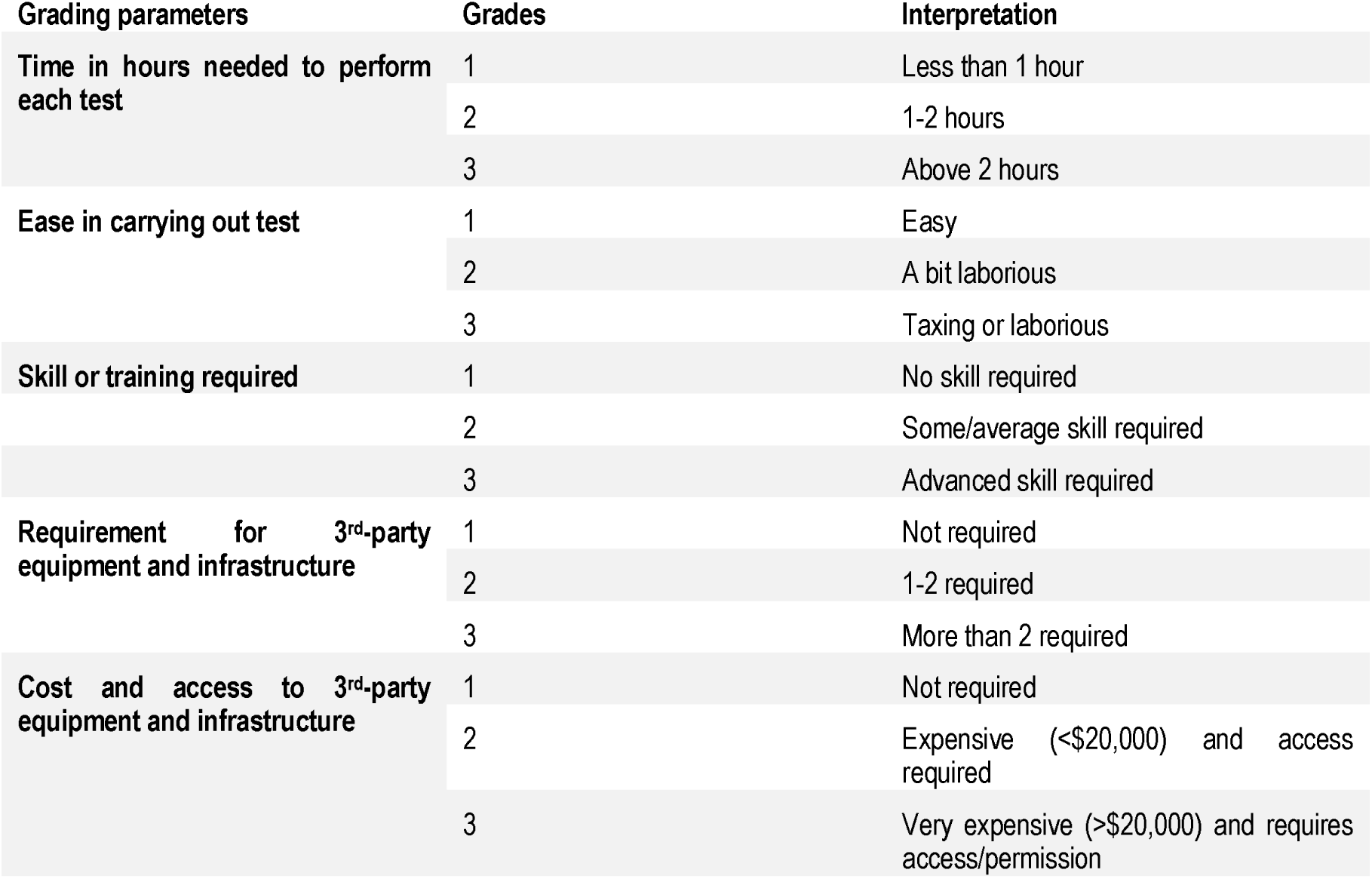
Grading criteria and interpretation for the colistin diagnostic tests.

## Results

The BMD identified 44 isolates as resistant to colistin and 40 as susceptible (Table 4). Only one isolate, an *E. coli*, was found to harbour the *mcr-1* gene per the PCR results (Table 4; Fig. 1). The isolates were identified by the MicroScan Walkaway to comprise of 12 species viz., *Enterobacter cloacae* (n=27), *Klebsiella pneumoniae* (n=12), *Salmonella enterica* (n=12), *Acinetobacter baumannii* (n=10), *Escherichia coli* (n=9), *Klebsiella oxytoca* (n=4), *Pseudomonas aeruginosa* (n=4), *Proteus mirabilis* (n=2), *Burkholderia cepacia* (n=1), *Hafnia alvei* (n=1), *Providencia stuartii* (n=1), and *Tatumella* spp. (n=1) (Figure 2A). CHROMagar COL-*APSE* supported the growth of 46 (55%) isolates, which were identified as COL-R (colistin-resistant) isolates with substantial false positives and false negatives (Tables 4-5; Figure 2B & 2C). The cultured isolates resulted in colony growth and identification of 22 isolates (26%) as COL-R coliforms, 15 isolates (18%) as COL-R *Acinetobacter* spp, three isolates (4%) as COL-R *Pseudomonas spp*, and six isolates (7%) as *E. coli*. A total of 38 isolates (45%) were not identified as COL-R by CHROMagar COL-*APSE*, most of which were susceptible (Tables 4-5). It is interesting to note that although this media suppresses the growth of susceptible strains, some grew on it, resulting in a sensitivity and specificity of 82.05% and 66.67% respectively. As well, the species prediction of CHROMagar COL-*APSE* mostly mirrored that of the MicroScan with some few disagreements (Tables 4-5). The CHROMagar COL-*APSE* was thus the least sensitive and least specific among all the tests (Table 5; Figure 3).

**Table 4.** General Table of results for all the four tests, multiplex PCR and BMD.

|  | BMD |  | CMT |  | ComASP™ |  | MicroScan |  |  | CHROMagar™ | Multiplex PCR results |
| --- | --- | --- | --- | --- | --- | --- | --- | --- | --- | --- | --- |
| Isolate | MIC | Int<sup>1</sup> | MIC | Int | MIC | Int | MIC | Int | ID | ID | *mcr-1* to *mcr-5* gene |
| **Amp1** | 32 | R | 2 | *mcr- neg*<sup>2</sup> | □ 16 | R | □ 4 | R | *K. pneumoniae* | COL-R<sup>3</sup> Coliform | Negative |
| **Amp2** | 32 | R | 4 | *mcr- neg* | □ 16 | R | □ 4 | R | *K. pneumoniae* | COL-R Coliform | Negative |
| **Amp3** | 8 | R | 1 | *mcr- neg* | 8 | R | □ 4 | R | *K. pneumoniae* | COL-R Coliform | Negative |
| **Amp4** | □ 64 | R | □ 64 | *mcr- neg* | □ 16 | R | □ 4 | R | *K. pneumoniae* | COL-R Acinetobacter | Negative |
| **Amp5** | 0,125 | S | 0,125 | *mcr- neg* | 0,25 | S | NR | R | *E. cloacae* | COL-R *E. coli* | Negative |
| **Amp6** | 0,25 | S | 0,125 | *mcr- neg* | 1 | S | NR | R | *E. cloacae* | COL-R *Pseudomonas* | Negative |
| **Amp7** | 0,5 | S | 0,125 | *mcr- neg* | 0,5 | S | NR | S | *A. baumannii* | COL-R coliform | Negative |
| **Amp8** | 0,5 | S | 0,5 | *mcr- neg* | 0,5 | S | NR | S | *A. baumannii* | COL-S<sup>4</sup> organism | Negative |
| **Amp9** | 0,5 | S | 0,5 | *mcr- neg* | 0,5 | S | NR | S | *A. baumannii* | COL-S organism | Negative |
| **Amp10** | 2 | S | 0,5 | *mcr- neg* | 2 | S | NR | S | *A. baumannii* | COL-S organism | Negative |
| **Amp11** | 2 | S | 0,25 | *mcr- neg* | 4 | R | □ 4 | R | *P. mirabilis* | COL-S organism | Negative |
| **Amp12** | 0,5 | S | 0,125 | *mcr- neg* | 2 | S | NR | R | *S. enterica* | COL-S organism | Faint bands suspected to be *mcr-2/3* |
| **Amp13** | 0,5 | S | 0,125 | *mcr- neg* | 0,5 | S | NR | R | *A. baumannii* | COL-R Acinetobacter | Negative |
| **Amp14** | 2 | S | 0,5 | *mcr- neg* | 4 | R | NR | S | *S. enterica* | COL-S organism | Faint bands suspected to be *mcr-2/3* |
| **Amp15** | 64 | R | 64 | *mcr- neg* | 0,25 | S | NR | R | *A. baumannii* | COL-S organism | Faint bands suspected to be *mcr-2/3* |
| **Amp16** | 64 | R | 32 | *mcr- neg* | □ 16 | R | NR | R | *P. marabilis* | COL-R Acinetobacter | Faint bands |

<sup>1</sup> Breakpoint interpretation
<sup>2</sup> *Mcr-1*-negative; this could also mean *mcr-2/-3/-4*-negative for isolates with DNA bands on the gel within the size of *mcr-2*,-3 & -4
<sup>3</sup> Colistin resistant
<sup>4</sup> Colistin susceptible

|  |  |  |  |  |  |  |  |  |  |  |  |
| --- | --- | --- | --- | --- | --- | --- | --- | --- | --- | --- | --- |
|  |  |  |  |  |  |  |  |  |  |  | suspected to be *mcr-2/3* |
| **Amp17** | □ 64 | R | 64 | *mcr- neg* | 8 | R | □ 4 | R | *E. coli* | COL-S organism | Negative |
| **Amp18** | 0,125 | S | 0,125 | *mcr- neg* | 4 | R | NR | R | *A. baumannii* | COL-S organism | Faint bands suspected to be *mcr-4/16* |
| **Amp19** | 2 | S | 0,5 | *mcr- neg* | 4 | R | NR | S | *S. enterica* | COL-S organism | Negative |
| **A7a** | 4 | R | 2 | *mcr- neg* | 4 | R | NR | S | *S. enterica* | COL-R Acinetobacter | Negative |
| **D5** | 32 | R | 16 | *mcr- neg* | □ 16 | R | □ 4 | R | *K. pneumoniae* | COL-R Coliform | Faint bands suspected to be *mcr-2/3* |
| **C9** | 8 | R | 4 | *mcr- neg* | 16 | R | □ 4 | R | *E. coli* | COL-R *E. coli* | Negative |
| **H7** | 4 | R | 0,5 | *mcr- neg* | 8 | R | NR | R | *B. cepacia complex* | COL-R Acinetobacter | Negative |
| **A6** | 32 | R | 4 | *mcr- neg* | 16 | R | □ 4 | R | *E. coli* | COL-R *E. coli* | Negative |
| **F7** | □ 64 | R | □ 64 | *mcr- neg* | □ 16 | R | NR | R | *E. cloacae* | COL-R Acinetobacter | Negative |
| **C1** | 4 | R | 0,25 | *mcr- neg* | 4 | R | NR | R | *S. enterica* | COL-R Coliform | Negative |
| **I7** | 0,25 | S | 8 | *mcr- neg* | □ 16 | R | NR | R | *E. cloacae* | COL-R Coliform | Negative |
| **G6** | □ 64 | R | □ 64 | *mcr- neg* | □ 16 | R | NR | R | *E. cloacae* | COL-R Coliform | Negative |
| **G7** | 2 | S | 0,125 | *mcr- neg* | 16 | R | NR | S | *E. cloacae* | COL-R Coliform | Negative |
| **D3** | 4 | R | 0,5 | *mcr- neg* | 2 | S | NR | R | *S. enterica* | COL-R Acinetobacter | Faint bands suspected to be *mcr-3/4* |
| **D2** | 4 | R | 2 | *mcr- neg* | □ 16 | R | NR | R | *S. enterica* | COL-R Acinetobacter | Negative |
| **G1** | □ 64 | R | □ 64 | *mcr- neg* | □ 16 | R | NR | S | *E. cloacae* | COL-R Coliform | Negative |
| **E8** | □ 64 | R | 8 | *mcr- neg* | □ 16 | R | NR | R | *E. cloacae* | COL-R Coliform | Negative |
| **D4** | 16 | R | 64 | *mcr- neg* | □ 16 | R | □ 4 | R | *K. pneumoniae* | COL-R Coliform | Faint bands suspected to be *mcr-2/3/4* |
| **B1** | □ 64 | R | □ 64 | *mcr- neg* | □ 16 | R | NR | R | *A. baumannii* | COL-R Coliform | Faint bands suspected to be *mcr-* |
| **A7b** | 4 | R | 4 | mcr- neg | □ 16 | R | □ 4 | R | *Hafnia alvei* | COL-R *E. coli* | Negative |
| **A8** | □ 64 | R | 16 | mcr- neg | □ 16 | R | NR | R | *E. cloacae* | COL-R Coliform | Negative |
| **B6** | □ 64 | R | 16 | mcr- neg | □ 16 | R | NR | R | *E. cloacae* | COL-R *Acinetobacter* | Negative |
| **C4** | 8 | R | 4 | mcr- neg | 4 | R | NR | R | *S. enterica* | COL-R *Acinetobacter* | Negative |
| **A5** | □ 64 | R | □ 64 | mcr- neg | □ 16 | R | NR | R | *Tatumella species* | COL-R Coliform | Faint bands suspected to be mcr-2/3 |
| **F6** | □ 64 | R | 64 | mcr- neg | □ 16 | R | NR | R | *E. cloacae* | COL-R Coliform | Negative |
| **D1a** | 8 | R | 4 | mcr- neg | 8 | R | NR | R | *E. cloacae* | COL-R *Pseudomonas* | Negative |
| **D1b** | 32 | R | 0,125 | mcr- pos | 4 | R | □ 4 | R | *E. coli* | COL-R *E. coli* | Mcr-1 |
| **D9** | 16 | R | 8 | mcr- neg | □ 16 | R | NR | R | *S. enterica* | COL-R Coliform | Negative |
| **S1** | 32 | R | 16 | mcr- neg | 16 | R | NR | R | *S. enterica* | COL-S organism | Negative |
| **S2** | 8 | R | 0,5 | mcr- neg | 16 | R | ≤2 | S | *Pseudomonas aeruginosa* | COL-R *Acinetobacter* | Negative |
| **S3** | 0,5 | S | 0,125 | mcr- neg | 16 | R | ≤2 | S | *Pseudomonas aeruginosa* | COL-R *Acinetobacter* | Negative |
| **S4** | 0,25 | S | 0,125 | mcr- neg | 0,5 | S | ≤2 | S | *Pseudomonas aeruginosa* | COL-R *Pseudomonas* | Negative |
| **S5** | 0,5 | S | 0,125 | mcr- neg | 0,5 | S | ≤2 | S | *Pseudomonas aeruginosa* | COL-R *Acinetobacter* | Negative |
| **S6** | 32 | R | 64 | mcr- neg | □ 16 | S | NR | S | *E. cloacae* | COL-S organism | Negative |
| **S7** | 0,5 | S | 0,125 | mcr- neg | 0,5 | S | NR | S | *E. cloacae* | COL-S organism | Negative |
| **S8** | 0,25 | S | 0,125 | mcr- neg | 8 | R | NR | S | *E. cloacae* | COL-S organism | Negative |
| **S9** | 0,125 | S | 0,125 | mcr- neg | 0,25 | S | ≤2 | S | *E. coli* | COL-S organism | Negative |
| **S10** | 0,125 | S | 0,125 | mcr- neg | 0,25 | S | ≤2 | S | *E. coli* | COL-S organism | Negative |
| **S11** | 0,5 | S | 0,125 | mcr- neg | 0,25 | S | 4 | R | *E. coli* | COL-S organism | Negative |
| **S12** | 0,125 | S | 0,125 | mcr- neg | 0,25 | S | ≤2 | S | *E. coli* | COL-R *E. coli* | Negative |
| **S13** | 0,125 | S | 1 | mcr- neg | 8 | R | ≤2 | S | *K. oxytoca* | COL-R Coliform | Negative |
| **S14** | 0,125 | S | 0,5 | mcr- neg | 0,25 | S | ≤2 | S | *K. oxytoca* | COL-S organism | Negative |
| **S15** | 1 | S | 0,25 | mcr- neg | 0,25 | S | ≤2 | S | *K. pneumoniae* | COL-R Coliform | Negative |
| **S16** | 0,125 | S | 0,125 | mcr- neg | 0,25 | S | ≤2 | S | *K. oxytoca* | COLS-organism | Negative |
| **S17** | 0,25 | S | 0,125 | mcr- neg | 0,25 | S | ≤2 | S | *K. oxytoca* | COL-S organism | Negative |
| **S18** | 0,25 | S | 0,125 | mcr- neg | 0,25 | S | 4 | R | *K. pneumoniae* | COL-R Coliform | Negative |
| **S19** | 32 | R | 64 | mcr- neg | 0,5 | S | ≤2 | S | *K. pneumoniae* | COL-S organism | Negative |
| **S20** | 64 | R | 16 | mcr- neg | 0,5 | S | ≤2 | S | *E. coli* | COL-S organism | Negative |
| **S21** | □ 64 | R | □ 64 | mcr- neg | □ 16 | R | □ 4 | R | *Providencia stuartii* | COL-S organism | Negative |
| **S22** | □ 64 | R | 16 | mcr- neg | □ 16 | R | NR | R | *A. baumannii* | COL-S organism | Negative |
| **S23** | 1 | S | 0,125 | mcr- neg | 0,5 | S | NR | S | *A. baumannii* | COL-S organism | Negative |
| **S24** | 0,125 | S | 0,125 | mcr- neg | 4 | R | NR | S | *E. cloacae* | COL-R Coliform | Negative |
| **S25** | 0,25 | S | 0,125 | mcr- neg | 0,5 | S | NR | S | *E. cloacae* | COL-S organism | Negative |
| **S26** | 0,5 | S | 0,125 | mcr- neg | 0,25 | S | NR | S | *E. cloacae* | COL-S organism | Negative |
| **S27** | 0,125 | S | 0,125 | mcr- neg | 16 | R | NR | S | *E. cloacae* | COL-R Coliform | Negative |
| **S28** | 1 | S | 0,125 | mcr- neg | 2 | S | NR | R | *E. cloacae* | COL-R Coliform | Negative |
| **S29** | 1 | S | 0,125 | mcr- neg | 0,25 | S | NR | S | *E. cloacae* | COL-S organism | Negative |
| **S30** | 0,5 | S | 0,125 | mcr- neg | 0,5 | S | NR | S | *E. cloacae* | COL-S organism | Negative |
| **S31** | 0,5 | S | 0,125 | mcr- neg | 1 | S | NR | R | *E. cloacae* | COL-S organism | Negative |
| **S32** | 2 | S | 0,25 | mcr- neg | 0,5 | S | NR | S | *E. cloacae* | COL-S organism | Negative |
| **S33** | 1 | S | 0,125 | mcr- neg | 8 | R | NR | R | *E. cloacae* | COL-S organism | Negative |
| **S34** | 0,25 | S | 0,125 | mcr- neg | 0,25 | S | ≤2 | S | *K. pneumoniae* | COL-S organism | Negative |
| **S35** | 1 | S | 0,125 | mcr- neg | 0,5 | S | □ 4 | R | *K. pneumoniae* | COL-S organism | Negative |
| **S36** | 0,5 | S | 0,125 | mcr- neg | 0,5 | S | ≤2 | S | *K. pneumoniae* | COL-S organism | Negative |
| **S37** | 1 | S | 0,25 | mcr- neg | 0,5 | S | NR | S | *E. cloacae* | COL-S organism | Negative |
| **S38** | 0,5 | S | 0,5 | mcr- neg | 2 | S | NR | S | *E. cloacae* | COL-S organism | Negative |
| **S39** | 8 | R | 1 | mcr- neg | 8 | R | NR | R | *S. enterica* | COL-R Acinetobacter | Negative |
| **S40** | 16 | R | 1 | mcr- neg | 2 | S | NR | R | *S. enterica* | COL-R Acinetobacter | Negative |

**Table 5.**
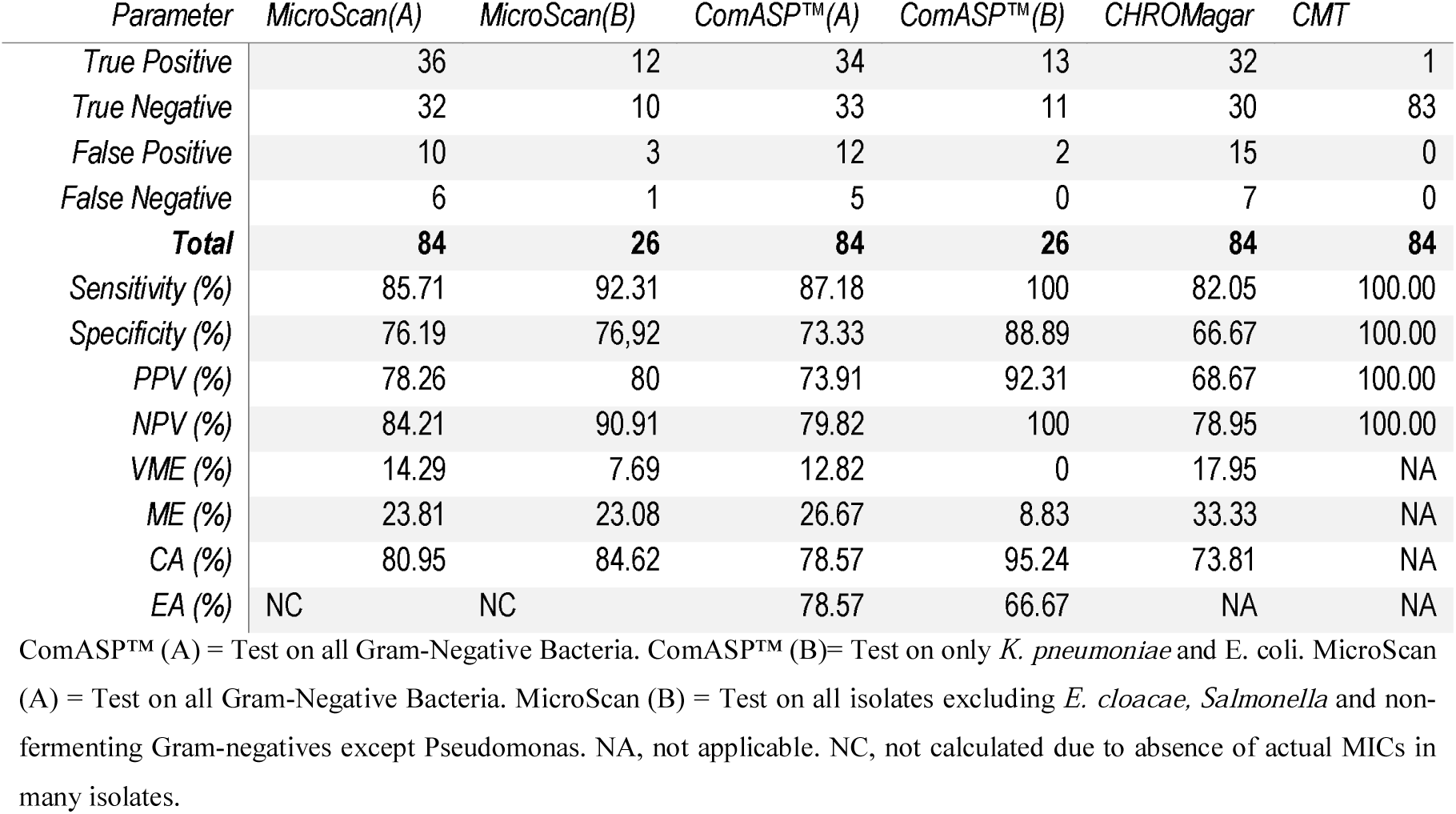
Diagnostic efficiency/performance of MicroScan, ComASP, and CHROMagar COL-APSE and CMT.

| Parameter | MicroScan(A) | MicroScan(B) | ComASP™(A) | ComASP™(B) | CHROMagar | CMT |
| --- | --- | --- | --- | --- | --- | --- |
| True Positive | 36 | 12 | 34 | 13 | 32 | 1 |
| True Negative | 32 | 10 | 33 | 11 | 30 | 83 |
| False Positive | 10 | 3 | 12 | 2 | 15 | 0 |
| False Negative | 6 | 1 | 5 | 0 | 7 | 0 |
| **Total** | **84** | **26** | **84** | **26** | **84** | **84** |
| Sensitivity (%) | 85.71 | 92.31 | 87.18 | 100 | 82.05 | 100.00 |
| Specificity (%) | 76.19 | 76.92 | 73.33 | 88.89 | 66.67 | 100.00 |
| PPV (%) | 78.26 | 80 | 73.91 | 92.31 | 68.67 | 100.00 |
| NPV (%) | 84.21 | 90.91 | 79.82 | 100 | 78.95 | 100.00 |
| VME (%) | 14.29 | 7.69 | 12.82 | 0 | 17.95 | NA |
| ME (%) | 23.81 | 23.08 | 26.67 | 8.83 | 33.33 | NA |
| CA (%) | 80.95 | 84.62 | 78.57 | 95.24 | 73.81 | NA |
| EA (%) | NC | NC | 78.57 | 66.67 | NA | NA |

ComASP™ (A) = Test on all Gram-Negative Bacteria. ComASP™ (B) = Test on only *K. pneumoniae* and *E. coli*. MicroScan (A) = Test on all Gram-Negative Bacteria. MicroScan (B) = Test on all isolates excluding *E. cloacae*, *Salmonella* and non-fermenting Gram-negatives except *Pseudomonas*. NA, not applicable. NC, not calculated due to absence of actual MICs in many isolates.

**Figure 1:**
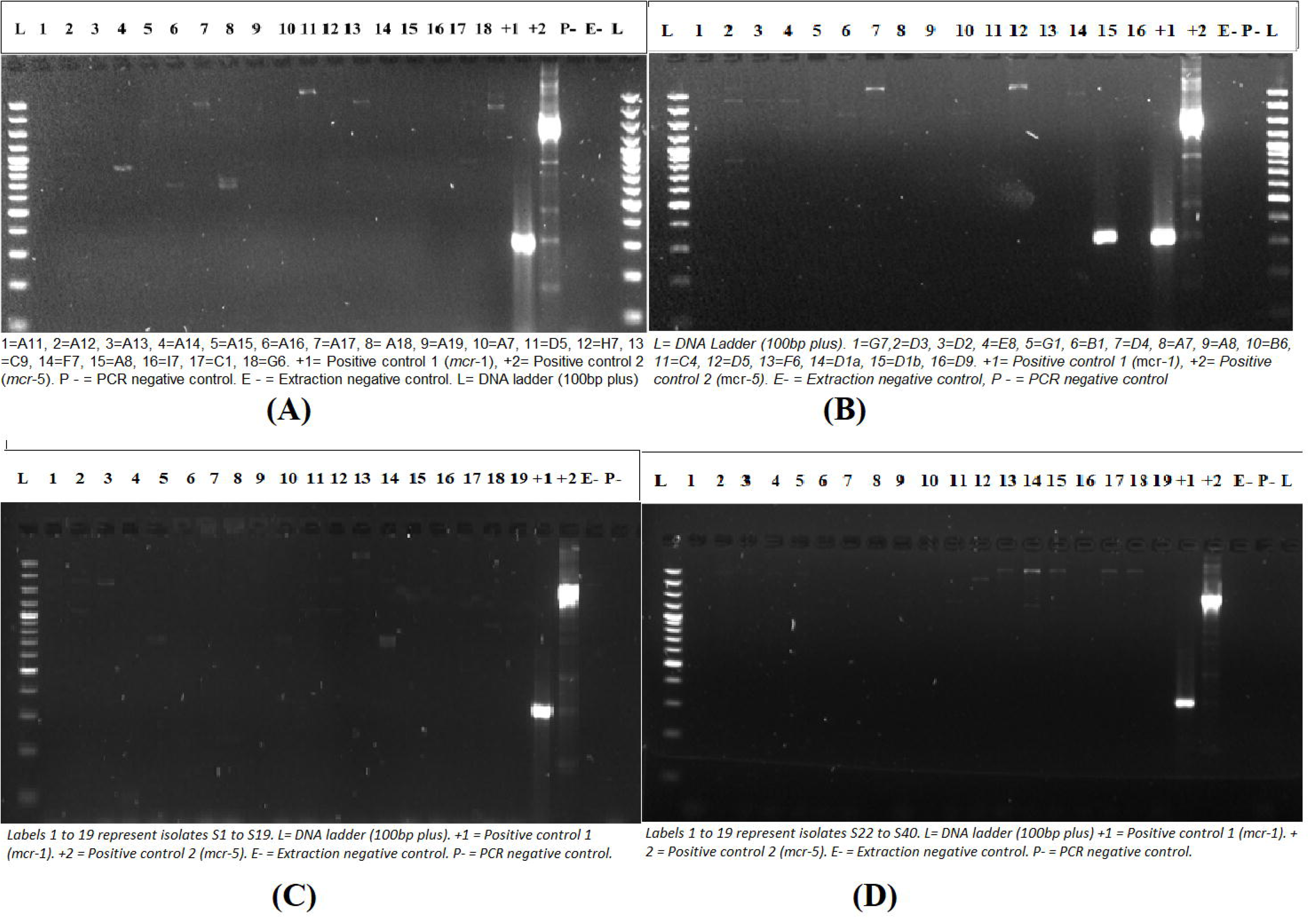
Gel electrophoresis run of *mcr-1* to *mcr-5* amplicons for all 84 samples. Image (A) contains results for 18 isolates, (B) contains results for 16 isolates, and (C) and (D) contains 19 isolates each. On the sides are the ladders (L) and controls. The *mcr-1-*positive isolate, D1b, is shown in (B).

**Figure 2:**
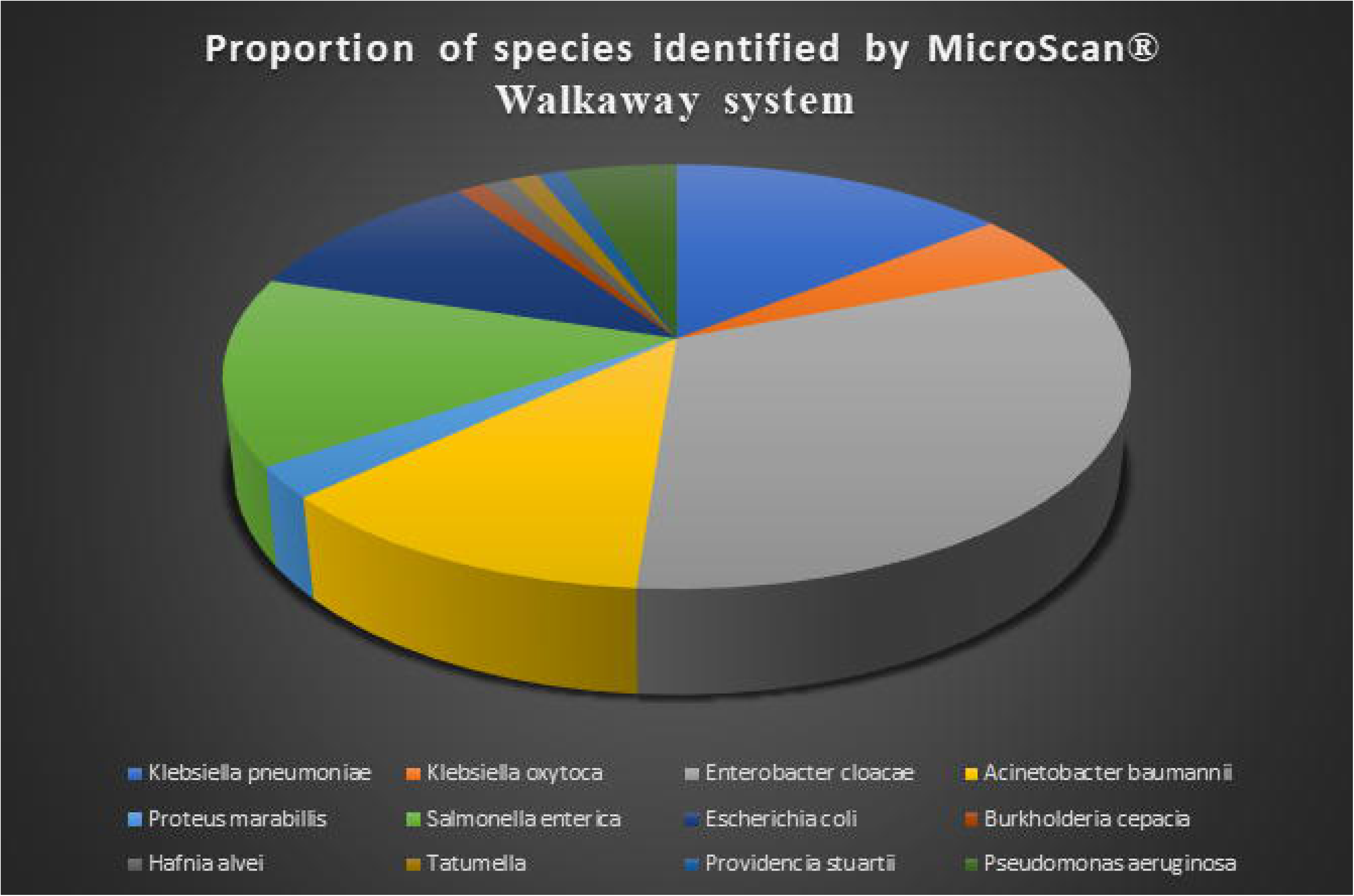
A pie chart showing the species distribution of the collected samples according to (A) MicroScan walkaway system results and (B) CHROMagar COL-APSE results. On (C) is shown the colony morphology on CHROMagar COL-APSE after overnight incubation at ambient air: (i) colistin-resistant *E. coli* are dark-pink to reddish, (ii) colistin-resistant coliforms are metallic blue, (iii) colistin-resistant *Pseudomonas* are translucent, +/- natural pigmentation cream to green, (iv) colistin-resistant *Acinetobacter* are cream(ish) and (v) other Gram-negative colistin-resistant species are colourless.

**Figure 3:**
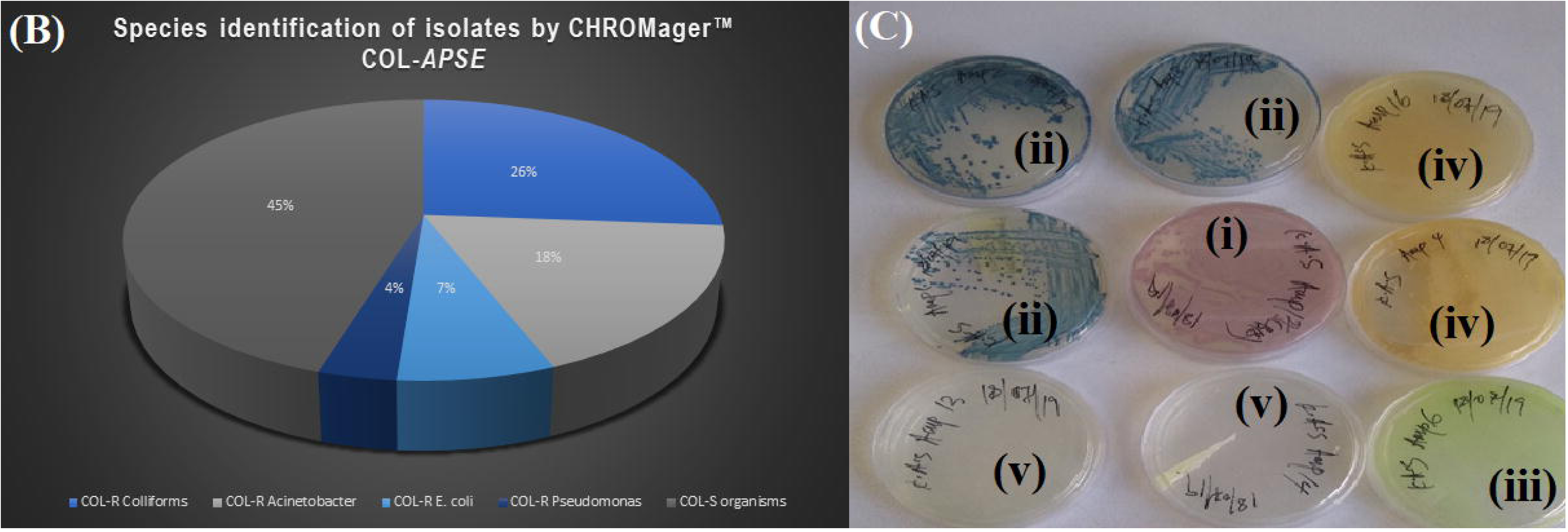
A graph illustrating the performance of the four colistin diagnostic tests. ComASP had the highest sensitivity, particularly for only *K. pneumoniae* and *E. coli*, followed by the MicroScan. For *mcr-1-*positive strains, the CMT was as efficient as the PCR. ComASP™ (A) = Test on all Gram-Negative Bacteria. ComASP™ (B)= Test on only K. pneumoniae and E. coli. MicroScan (A)= Test on all Gram-Negative Bacteria. MicroScan (B)= Test on all isolates excluding E. cloacae, Salmonella and non-fermenting gram-negatives except Pseudomonas.

However, the ComASP had an overall sensitivity of 87.18% and specificity of 73.33%, which respectively increases to 100% and 88.89%, when only *K. pneumoniae* and *E. coli* results are used (Table 5). The evaluable essential agreement was calculated by evaluating ComASP results (≤ 0.25 µg/mL or C 16 µg/mL) with BMD results (≤ 0.5µg/mL or≥ 16 µg/mL). The overall evaluable EA was 53%, but with the focus on *K. pneumoniae* and *E. coli* only, the evaluable EA was 66.67%. A PPV and NPV of 73.91% and 86.84% were recorded respectively, with a VME of 12.82% and a ME of 26.67%. ComASP had an EA of 78.57%.

Thus, the ComASP overall results (A, see Figure 3) proved quite more sensitive but less specific than the MicroScan system, which had an overall sensitivity and specificity of 85.71% and 76.19%. When *E. cloacae, Salmonella* and non-fermenting Gram-negatives except Pseudomonas spp. are excluded from the MicroScan results, its sensitivity, but not specificity, increased substantially to 92.31% (Table 5). The CMT was able to identify the only *mcr-1* positive isolate and the 83 non-*mcr-*positive isolates, resulting in a sensitivity and specificity of 100% (Table 5). Further, reductions in MICs (n=54), and in some cases increments (n=5 isolates), were observed after adding DPA to the medium. Among those with reductions or increments, 11 had visible bands on the gel, one of which was the *E. coli* (D1b) isolate that was *mcr-1* positive (Figures 1A & B; Table 4).

Notably, all of these 10 isolates with bands on the gel were classified as *mcr-*negative per the interpretation criteria of the CMT ^9,15^. However, there were conflicting MICs between the BMD, ComASP and MicroScan for these isolates: Amp16 (*P. mirabilis*), D5 (*K. pneumoniae*), C9 (*E. coli*), G6 (*E. cloacae*), D4 (*K. pneumoniae*) and A5 (*Tatumella* spp.) were all resistant by the BMD, ComASP Colistin and MicroScan; Amp12 (*S. enterica*) was susceptible to all the tests but was colistin resistant per the BMD; Amp14 (*S. enterica*) was colistin resistant per the ComASP Colistin, but susceptible per the BMD and MicroScan; Amp15 (*A. baumannii*) was resistant per the BMD and MicroScan, but susceptible per ComASP Colistin; Amp18 (*A. baumannii*) and D5 (*K. pneumoniae*) were both resistant per ComASP and MicroScan, but susceptible per BMD. Thus, most of the isolates with bands were colistin resistant per the BMD (n=6).

In terms of cost per sample, the MicroScan was the most expensive at a cost of R221.6 ($15.0), followed by CHROMagar COL-*APSE* (R118.3; $8.0), M-PCR (R75.1; $5.1), CMT (R20.1; $1.4) and ComASP Colistin (R2.64; $0.2). With regards to time and ease of application, the CHROMagar was the easiest to perform, followed by ComASP Colistin, M-PCR, MicroScan, CMT and BMD. However, the MicroScan and PCR required 3rd-party equipment and advanced skill that could be a challenge for lower income countries or under-resourced laboratories. With the exception of M-PCR, all the tests had a TAT (turnaround time) of 18-24 hours (Table 6).

**Table 6.** Diagnostic simplicity, time, required skill, equipment and ease-of-application grading for the evaluated tests: MicroScan, ComASP, CHROMagar COL-APSE, CMT, M-PCR and BMD.

| Test criteria | Grade |  |  |  |  |  |
| --- | --- | --- | --- | --- | --- | --- |
|  | CHROMagar COL-APSE (TAT = 18-24 hrs) | ComASP (TAT = 18-24 hrs) | M-PCR (TAT = 1-3 hrs) | MicroScan (TAT* = 18-24 hrs) | CMT (TAT = 18-24 hrs) | BMD (TAT = 18-24 hrs) |
| Time in hours needed to perform each test | 1 | 1 | 2 | 2 | 3 | 3 |
| Ease in carrying out test | 1 | 2 | 2 | 2 | 3 | 3 |
| Required training or skill | 1 | 1 | 3 | 3 | 2 | 2 |
| Requirement for 3<sup>rd</sup>-party equipment and/or infrastructure | 2 | 2 | 2 | 2 | 2 | 2 |
| Cost and access to 3<sup>rd</sup>-party equipment and/or infrastructure | 1 | 1 | 2 | 3 | 1 | 1 |
| Total grading | **6** | **7** | **11** | **12** | **11** | **11** |

\*TAT: turnaround time, M-PCR: multiplex PCR, CMT: colistin MAC Test, BMD: broth microdilution

## Discussion

The importance of colistin in the treatment of fatal infections caused by carbapenem-resistant GNB makes the search for an efficient colistin diagnostic an essential and life-saving exercise. However, finding a diagnostic tool that can be easily adapted into the workflow of routine microbiology laboratories remains a challenge as the recommended colistin sensitivity testing method, the BMD, is labour intensive ^9^. Moreover, disc diffusion and E-test, which are simpler to perform, are fraught with inherent errors due to difficulty in colistin diffusion through agar ^9^. Several rapid and non-rapid assays have been designed and proposed to help detect colistin directly from clinical samples or from culture, yet challenges still exist in terms of diagnostic performance, TAT, required skill, sample-processing cost etc. ^9^ In this study, selected culture-based diagnostics were evaluated to determine their relative diagnostic efficiencies viz., sensitivities, specificities, NPV, PPV, ME, VME, EA and CA, required skill, ease of application, requirement for 3^rd^-party equipment/infrastructure and sample-processing cost to advise clinical and diagnostic laboratories, particularly in resource-constrained settings.

ComASP Colistin is a BMD-based method used to determine colistin MICs (Carreto *et al*., 2018)^16^. The 96-well BMD method is a very labour-intensive technique that requires a highly skilled professional, all plates and antibiotic concentrations must be manually prepared, and it is very time-consuming (Table 6) ^9^. In this study, ComASP™ was able to successfully give colistin MICs for all tested isolates, with an essential agreement of 78.57%. The assay recorded a CA of 79.76% and a VME of 12.82% (lowest out of all the evaluated tests) and a ME of 26.67%. The VME and ME are high and unacceptable, but this can be explained by the fact that this assay was challenged with Gram-negative bacteria, which include the non-fermenters, compared to other studies where the assay was only used to investigate colistin MICs for *E. coli* and *K. pneumoniae* (Table 7) ^17^.

**Table 7.** Comparison of performance of ComASP™ colistin in our study with other studies only focusing on K. pneumoniae and E. coli isolates.

|  | (Matuschek et al., 2018)<sup>18</sup> | (Carreto et al., 2018)<sup>16</sup> | (Galani et al., 2018)<sup>17</sup> | This study |
| --- | --- | --- | --- | --- |
|  | *E. coli* & *K. pneumoniae* (n=32, 19COL-R) | *K. pneumoniae* (n=76, 23COL-R) | *K. pneumoniae* (n=392, 141COL-R) | *E. coli* & *K. pneumoniae* (n=21, 12COL-R) |
| EA | 96% | 94.7% | 94.9% | 66.67% |
| CA | 94% | 98.7% | 97.2% | 95.92% |
| MEs | 15.4% | 1.9% | 3.98% | 8.83% |
| VMEs | 0 | 0 | 0.7% | 0 |

EA: essential agreement, CA: categorical agreement, MEs: major error, VME: very major error. Table 6 above indicates ComASP™ results from other studies compared to results in this study. In this study, the CA, MEs and VMEs for ComASP™ Colistin were consistent with other studies but recorded the lowest EA.

Thus, upon excluding results from the non-fermenting Gram-negative bacteria and using only *K. pneumoniae* and *E. coli* as done in previous works ^16,18^, the ComASP performed way better, with a sensitivity and specificity of 100% and 88.89% respectively (Table 5; Figure 3). A PPV and NPV of 92.31% and 100% were respectively recorded whilst a CA, EA and ME of 95.24%, 66.67% and 8.83% were also respectively obtained; no VMEs were observed (Table 7). The EA could only be calculated by evaluating ComASP results (≤ 0.25 µg/mL or C 16 µg/mL) with BMD results (≤ 0.5µg/mL or≥ 16 µg/mL), given the differences between the colistin concentrations between the ComASP and the BMD. Thus, ComASP is best suited for detecting COL-R *K. pneumoniae* and *E coli* isolates as observed in a previous study by Matuschek and coworkers (2018) who concluded that the ComASP™ can reliably identify most COL-R K. *pneumoniae* and *E. coli* isolates ^18^. ComASP Colistin is easy to perform and interpret the results. It is not too costly and can be easily used in resource poor laboratories.

Overall, the ComASP (formerly SensiTest Colistin) performed better than the other diagnostics, followed closely by the MicroScan (Figure 3). Notably, these two high-performing tests recorded higher diagnostic performances when certain species that lowered the tests’ sensitivities were excluded, suggesting that not all species can be efficiently analysed with these two tests. Specifically, the MicroScan also increased in sensitivity after excluding *E. cloacae, Salmonella* and Gram-negative non-fermenters except *Pseudomonas* spp. Therefore, results on species that performed poorly on these tests should be further verified with other tests that are efficient in detecting colistin resistance in those species to avoid making fatal diagnostic or therapeutic decisions. For instance, laboratories that are already using the MicroScan can augment it with the ComASP. In terms of cost, ease of application and required skill, the ComASP further had an advantage over the MicroScan, although it should be noted that the MicroScan can determine the AST of several antibiotics whilst the ComASP is only designed for colistin ^9^.

The MicroScan system is laborious due to the daily maintenance of the instrument required and barcoding of the panels. The system is very costly as there are no panels available for only colistin AST. The system also requires highly skilled or trained personnel to operate it (Tables 2 & 5). It is important to take note of factors stated by Jayol and co-workers (2018)^19^ that the MicroScan system’s procedural manual indicates that results for *E. cloacae, Salmonella* and non-fermenting Gram-negative bacteria, except *Pseudomonas* spp., must not be reported. The latter is a major challenge and, in this study, all results for our *E. cloacae, Salmonella* and non-fermenting Gram-negative bacteria did not give MIC values as they returned as “Non-readable result” (Table 3). Hence, we only regarded the MicroScan’s results without the *E. cloacae, Salmonella* and non-fermenting Gram-negative bacteria except *Pseudomonas* as suitable to evaluate with the other tests. Consequently, the MicroScan was able to detect 92.31% of the isolates that were resistant to colistin by the gold standard. The MicroScan had a low specificity of 76.92% after the ComASP Colistin’s 88.89 (B; see Figure 3). This is further supported by a study in South Africa by Mohlabeng and colleagues (2017), where the MicroScan’s efficacy in reporting Carbapenemase-Producing *Enterobacteriaceae* was investigated and it was concluded that the MicroScan Walkaway system is sensitive but lacks specificity ^20^. One isolate was falsely detected as susceptible, explaining the 7.69% VME rate.

CHROMagar COL-*APSE* is a selective culture media used to isolate and differentiate colistin-resistant *Acinetobacter, Pseudomonas, Stenotrophomonas* and *Enterobacteriaceae* species with intrinsic, acquired or novel mechanisms of COL-resistance ^21^. CHROMagar identified and differentiated 46 isolates as colistin resistant, with majority being coliforms, *Acinetobacter* and *Pseudomonas spp*. (Figure 2B-C). Interestingly, 7% of the colistin-resistant isolates were identified to the species level as *E. coli*, including the *mcr-*1 positive *E. coli* isolate, which further supports CHROMagar COL-*APSE*’s ability to recover colistin-resistant isolates with acquired colistin resistance as stated by the developers of the medium ^21^. In this study, the CHROMagar had the highest VME and ME, which should be investigated further.

In a previous study, a 100% specificity and sensitivity was recorded for CHROMagar ^21^ whilst we recorded a specificity of 66.67% and a sensitivity of 82.05%. This discrepancy could be due to the other study’s use of serial dilutions in broth whilst we evaluated the performance of the media using cultured bacteria; this might be a limitation in our study that can be further investigated. A recent study by Thiry *et al*. (2019) evaluated CHROMagar COL-*APSE* against CHROMID Colistin_R and agar disk diffusion and concluded that growth on the media corelates with resistant results on the disk diffusion and not with the presence of the *mcr* genes ^10^. Our study supports this as growth on the CHROMagar in this study was observed for most isolates that did have any *mcr* genes in the PCR assay; however, this study had only one *mcr-*positive isolate, limiting a comprehensive comparison and conclusion. Nevertheless, these findings indicate that the mechanism of resistance does not limit the ability of CHROMagar to isolate and differentiate colistin-resistant organisms. CHROMagar COL-*APSE* plates are easy to prepare and interpret the results. Moreover, following the manufacturer’s instructions is easy, and the technique does not require a skilled individual (Tables 2 & 5). The medium is cost effective and only requires overnight incubation until results can be obtained.

The PCR assay detected several bands, which were within the size specifications of *mcr-2/-3/-4* genes (Figure 1): visible bands were seen in Fig. 1A (lanes 4, 6, 7, 8, 13) and 1B (lane 2) between the *mcr-1* and *mcr-5* positive control bands, which we strongly suspect could be *mcr-2, mcr-3* and *mcr-4;* nevertheless, we were unable to confirm these with sequencing and controls for these genes. However, a single *E. coli* isolate (D1b) was found to be *mcr*-1 positive by the multiplex PCR as shown in Figure 1. The latter is not surprising as previous works in South Africa has shown the *mcr*-1 gene to be dominant in the country and has been isolated in Johannesburg, Pretoria and Cape Town from *E. coli* isolates ^22^. Isolates D3 (Fig. 1B lane) and B1 showed non-specific bands around the *mcr*-3 and *mcr*-4 genes respectively, but we could not confirm these further due to restricted funding. Indeed, the other faint bands on the gels could be other *mcr* variants beyond *mcr-1* to *mcr-5* as *mcr-9*.*1* was recently reported from the same laboratory ^23^. Plans are far advanced to investigate these and add more *mcr* variants in future studies. The multiplex PCR is highly sensitive and specific, but it could be confirmed through whole-genome sequencing (WGS) to ascertain the presence or absence of other *mcr*-genes and alleles. The PCR is expensive, quite time-consuming and requires a highly skilled individual to perform and analyse the results (Tables 2 & 5).

We included the multiplex PCR assay in this evaluation to enable us compare its cost, skill, TAT, and efficiency in detecting *mcr-mediated* colistin resistance compared to non-molecular tests in resource-constrained settings. It was also necessary to determine the colistin resistance mechanisms underlying the observed resistance phenotypes in order to properly interpret the data of the various diagnostics results. It is worthy to note that not all *mcr-* positive strains express colistin resistance but can be expressed in the presence of a suitable promoter ^24^. Such incidences are a major disadvantage of phenotypic tests that report sensitive results for clinical samples, but patients fail to respond to therapeutics to which the strains were found susceptible *in-vitro*. This gives molecular tests a major advantage over non-molecular tests.

The CMT is a DPA chelator-based tool used to detect the presence of *mcr* genes ^9^. Herein, no ≥8 fold-increases in MICs were observed for all the *mcr*-negative isolates, consistent with a study by Coppi and colleagues (2018) where their colistin MICs neither changed nor increased or were at most reduced by two folds for 13 *mcr*-negative colistin-resistant strains. The isolates for which amplicon bands were observed within the *mcr-2/-3/-4* size regions were mostly colistin resistant per the BMD, MicroScan and/or ComASP colistin tests, but could not be detected by the CMT test although they had decreased MICs in the presence of DPA. This could suggest that the CMT is most efficient in detecting *mcr-1* and not other *mcr* variants or that a revision in the MIC cut-off differential for other *mcr* variants could enable it identify them perfectly. Nevertheless, the observed MIC reductions in the presence of DPA does suggest the presence of a metallo-enzyme-mediated colistin resistance in the isolates, which we hope to investigate further in future studies.

Notably, Coppi et al. (2018) observed that the CMT was not efficient in *K. pneumoniae* and we could not confirm this due to the absence of an *mcr-1-*positive *K. pneumoniae* isolate. Furthermore, the ability of the CMT to detect other *mcr* variants could not be established herein due to the inability of CMT to confirm the presence of other *mcr* variants observed on the gels. These are limitations to this study, although plans are far advanced to include other control *mcr* variants in subsequent evaluations. Our *mcr*-1 positive *E. coli* isolate had an 8-fold MIC reduction, similar to that reported by Coppi et al. (2018) wherein their 59 *mcr*-1-positive *E. coli* strains had a ≥ 8-fold reduction in colistin MICs in the presence of DPA. Unlike the other tests, the CMT assay cannot detect colistin-resistant isolates that are *mcr-*negative and can be a major limitation in settings where *mcr* genes are absent. Thus, coupled with its laborious and time-consuming nature, it might be difficult for the CMT to be adopted in clinical microbiology laboratories. However, where access to PCR is a challenge, it can be a cheaper, albeit time-consuming, alternative.

The presence of only a single well-defined *mcr-1-*positve isolate is a major limitation for this study as a single isolate cannot be used to make strong conclusions based on statistical inference. However, it is worth noting that *mcr-1* remains the most clinically and epidemiologically important *mcr* variant in all countries worldwide, making its inclusion very critical and important. Hence, the absence of the other less clinically important *mcr* variants cannot substantially affect the importance of this work in terms of its application in other settings. As well, we could not include other rapid screening and biochemical tests such as the SuperPolymyxin, LBJMR, the rapid Polymyxin NP test (RPNP) and commercial RPNP tests ^9^ due to logistical challenges.

## Conclusion

In this study, the ComASP was found to be the best-performing MIC determiner because it had the highest sensitivity and specificity with minimal errors but were more costly than the BMD. ComASP performed better than other alternative MIC determiners when only E. coli and *K. pneumoniae* are included in the analysis. If all other species are considered, ComASP performance is comparable to other alternatives with marginal differences. It is easy to use and can be employed in diagnostic laboratories that seek to conduct colistin susceptibility screening. The assay can be easily used in less-resourced laboratories without the need of a skilled personnel; the only requirement is an appropriate incubator. The MicroScan walkaway system is a very expensive tool that demands highly skilled personnel and is time-consuming. The MicroScan walkaway system was the second-best diagnostic with regards to its sensitivity and had the second lowest VMEs and lowest MEs, albeit it was the most expensive. It is also not ideal for colistin AST of non-fermenting Gram-negative bacteria and the BMD must be used to confirm the MicroScan’s colistin AST results for non-fermenters.

CHROMagar COL-*APSE* is more costly than the gold standard but can be used in any routine laboratory that has an incubator. However, its diagnostic performance was relatively poor, making it unsuitable for routine use in a diagnostic laboratory. The multiplex PCR is a sensitive and specific tool for detecting plasmid-mediated colistin resistance *mcr* genes, albeit it is expensive (fourth most expensive in this study), requires high skill and cannot be used to guide decisions on therapy. It is however ideal for surveillance in diagnostic laboratories without WGS capabilities for confirming the presence or absence of *mcr* genes in colistin-resistant isolates. The CMT is cost effective for detecting *mcr*-1-positive isolates (except in *K. pneumoniae*) in a setting without molecular assays such as the multiplex PCR or WGS; however, more work is needed to challenge this technique with different *mcr* positive strains including novel alleles. It’s efficacy in detecting *mcr* genes among *K. pneumoniae* also needs further investigation. Further studies must look at WGS and its performance in rapidly detecting *mcr-*harbouring strains of Gram-negative bacteria that are colistin-resistant^25^

### Ethical consent

This study is part of an umbrella study with the ethics no 483/2016. Ethical amendment for this study was applied for from the Research Ethics Committee of the Faculty of Health Sciences, University of Pretoria and was approved (ethics reference number 240/2019). Specimens were received and processed at the laboratories of the Department of Medical Microbiology, Faculty of Health Sciences, University of Pretoria. For the protection of patients from whom the specimens were taken, a numerical labelling system was developed to ensure that the identity of the patients remains undisclosed.

## Data Availability

All data are included in this article

## Acknowledgements

We acknowledge the kind support of Staff at the NHLS laboratory for their kind support. We are also exceptionally grateful to Kathy-Anne Strydom of Ampath Laboratories, Johannesburg, for donating to us some colistin-resistant strains for the evaluation.

## Transparency declaration

None

## Author contributions

*Study design*: JOS; *Supervision:* NMM & JOS; Laboratory work: AKS; Data analyses: AKS & JOS; *Manuscript write-up, tabulations, figures*: JOS

---

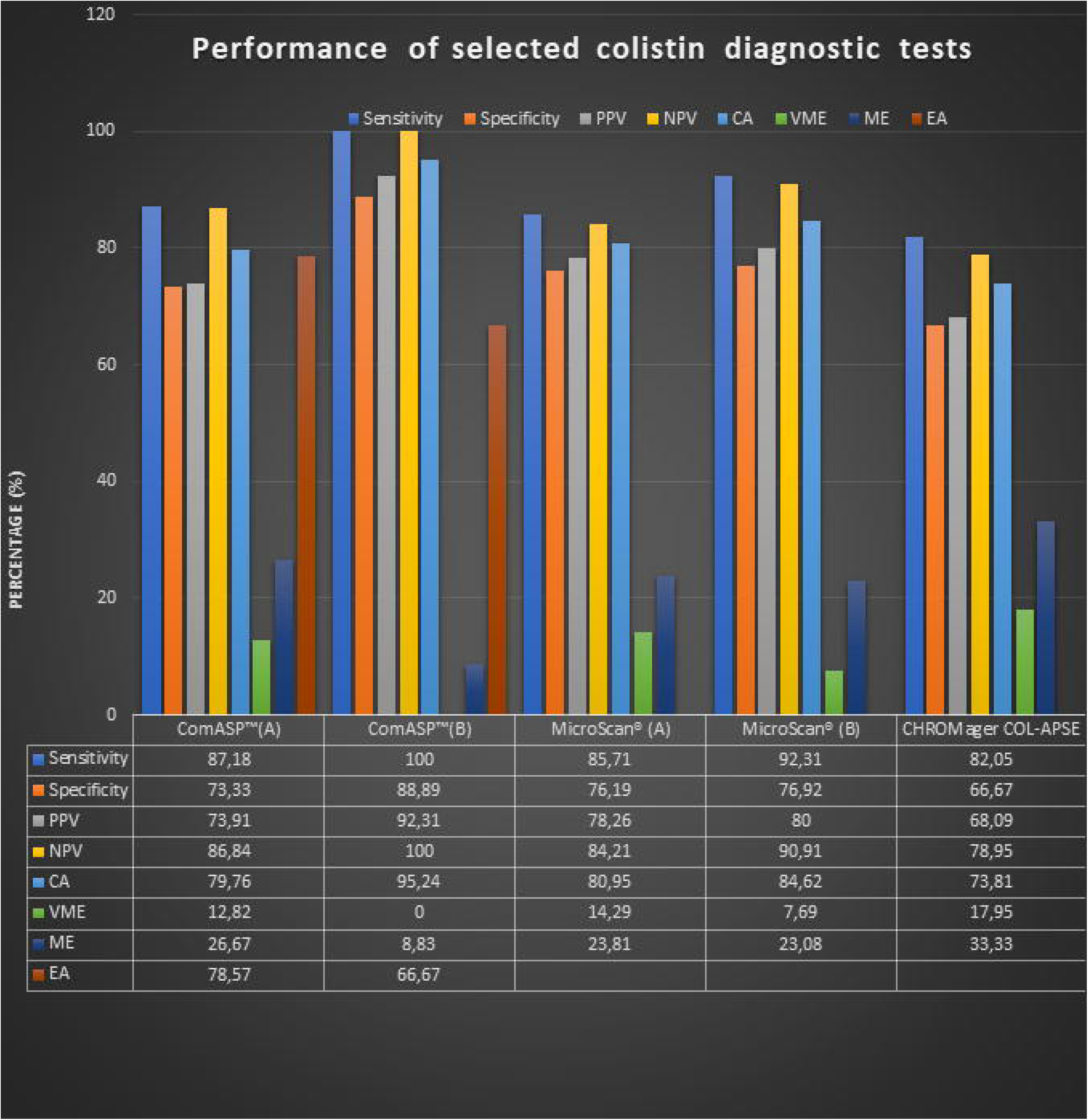

